# Persistence of respiratory, enteric, and fecal indicator viruses in fecal sludge from on-site sanitation in Dakar, Senegal

**DOI:** 10.1101/2024.08.07.24302194

**Authors:** Lorelay Mendoza Grijalva, Alsane Seck, Laura Roldan-Hernandez, Katherine E. Graham, Alexandria B. Boehm, William A. Tarpeh

## Abstract

As wastewater-based epidemiology (WBE) broadens its focus to include prevalent diseases with significant global health impact, existing surveillance systems concentrate on sewer-based infrastructure, which excludes the 2.7 billion people using non-sewered systems. To address this gap, our study explores the potential of fecal sludge treatment plants (FSTPs) for WBE, emphasizing the stability of virus RNA targets within pooled fecal sludge. We screened fecal sludge from a centralized treatment facility in Dakar, Senegal for SARS-CoV-2, human norovirus (HuNoV), and microbial source trackers (MSTs) pepper mild mottle virus (PMMoV) and tomato brown rugose fruit virus (ToBRFV). Decay kinetics of genomic RNA markers from these viruses were examined at 4 °C, 15 °C, and 30 °C over 70 days. Results indicate high persistence of viral targets in fecal sludge (T90 value of 3.3 months for exogenous SARS-CoV-2 N1 and N2, 6.2 months for ToBRFV), with all targets detected throughout the 70-day experiment under various temperatures with limited decay (<1 log10 reduction). This study addresses a crucial gap in understanding virus persistence in on-site sanitation systems by providing essential decay rate constants for effective target detection. Our results indicate that sampling at centralized facilities treating fecal sludge from on-site sanitation could facilitate localized pathogen surveillance in low-income settings.

**Highlights:**

- Investigation of the persistence of SARS-CoV-2, HuNoV, PMMoV, and ToBRFV genomic RNA in pooled fecal sludge derived from on-site sanitation systems.
- Novel microbial source tracker (MST), ToBRFV, exhibited comparable abundance to PMMoV, a well-established MST, in fecal sludge.
- No significant decay observed for HuNoV and PMMoV over 70 days at all temperature conditions (4, 15, and 30 °C).
- SARS-CoV-2 N1 and N2 showed T_90_ values of 3.3 months at 30 °C.
- Fecal sludge treatment plants offer a centralized sampling location for wastewater-based epidemiology, providing a strategic approach for monitoring public health.

**Graphical Abstract:** 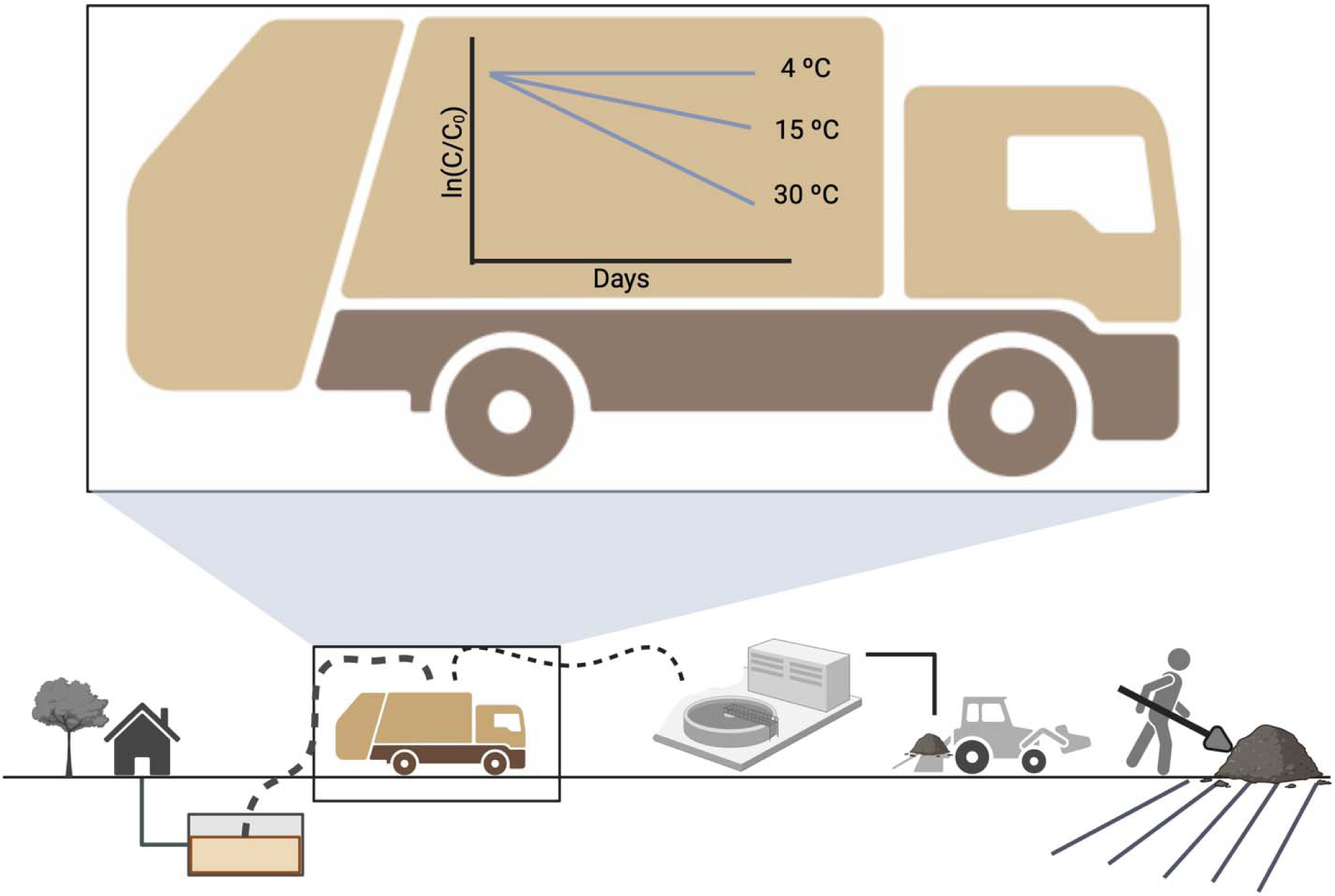

## Introduction

Wastewater-based epidemiology (WBE) emerged as a vital tool amid the COVID-19 pandemic, offering actionable public health insights that supplement traditional disease surveillance. Its pandemic response contributions include capturing asymptomatic carriers, identifying hotspots requiring resource deployment (Kirby *et al*., 2021; Diamond *et al*., 2022), and illustrating the regional spread of emerging variants (Izquierdo-Lara *et al*., 2021; Boehm *et al*., 2022). WBE has since extended its scope to the surveillance of other infectious diseases caused by viruses (CDC, 2022; Wolfe *et al*., 2023). While advancements in WBE have focused on urban, high-income areas served by centralized, sewered treatment facilities thus far (Naughton et al., 2021), on-site sanitation technologies, such as septic tanks and pit latrines, predominant in Sub-Saharan Africa (72%), Central Asia (68%), and Latin America and the Caribbean (31%) (Greene et al., 2021), are often overlooked. Despite serving 2.7 billion individuals, the potential to provide community disease surveillance for populations reliant on non-sewered, on-site sanitation systems has been highlighted (Capone et al. 2020; Li et al. 2023) but remains underexplored.

Conducting community health surveillance in non-sewered settings using fecal sludge presents context-specific challenges, such as scale, representativeness, and the stability of viral targets in on-site sanitation. Scale concerns arise because septic tanks and pit latrines typically serve individual households, making community surveillance resource-intensive. Attaining representativeness poses challenges in areas with limited sanitation infrastructure. Although environmental sampling can detect pathogens, the samples collected may generally lack information about the origin of flow inputs, the contributing population, and the environmental conditions between excretion and sampling. These inconsistencies may adversely impact the quality of measurements, thus challenging reproducibility and diminishing the ability to compare results between sites or studies. The limited understanding of target stability also increases the challenge of interpreting the data because on-site sanitation technologies may retain excreta for extended periods (i.e., months to years) before disposal (Jensen *et al*., 2009; Kumwenda, 2019; Musaazi *et al*., 2023).

Sampling from fecal sludge treatment plants (FSTPs) can begin to address these challenges; FSTPs serve as centralized facilities that collect and treat fecal sludge from diverse areas, and are common in urban and peri-urban areas (Atitaya Panuvatvanich, 2013; Sanitation Matters, 2013; Harada, Strande and Fujii, 2015). FSTPs receive sludge from trucks servicing on-site systems, offering a pooled sample more representative than individual sanitation systems. Factors like sewershed size, input frequency, and sources may be more feasibly estimated than from surface water or soil samples, offering clearer understanding into the environmental conditions affecting the integrity of the sampled wastewater. Leveraging existing waste operations in non-sewered localities like Dakar, Senegal, presents promising opportunities to survey community health. Compared to on-site sanitation systems alone, FSTPs serve a larger population (scale) and encompass a known or consistent group of people (representative); however, insights into the stability of viral targets within the community’s waste stream are unknown.

The stability of targets has been highlighted as an important research area within WBE (Capone et al., 2021; Li et al., 2021), and determining the persistence of viral RNA in fecal sludge from FSTPs is a sensible first step towards developing effective WBE programs in communities that use on-site sanitation systems. While numerous studies have explored virus decay in wastewater from sewered settings (Ahmed *et al*., 2020; Bivins *et al*., 2020; Fu *et al*., 2022; Roldan-Hernandez *et al*., 2022), research on pathogen persistence within on-site sanitation systems is limited. On-site systems, dealing with smaller volumes and potentially higher solids content, may not consistently receive household greywater. Their characteristics can differ from sewered wastewater, and wastewater composition is known to impact pathogen decay (Koné *et al*., 2007; Mehl *et al*., 2011; Darimani *et al*., 2015). Assessing their decay in context-specific matrices, such as fecal sludge from aggregated on-site sanitation systems, is vital due to potential variations in wastewater composition. Relying on decay values from other wastewater sources may not represent the unique characteristics of these sanitation contexts. Determining the decay rate of viral targets in fecal sludge may also offer insight into the maximum duration of sludge residence in on-site systems that still allow for their detection using molecular methods.

In this study, we sought to assess the feasibility of utilizing FSTPs to implement WBE. We focused on the stability of various viral genomic RNA targets in fecal sludge samples from a centralized fecal sludge treatment facility in Dakar. We screened for key respiratory, enteric, and fecal indicator viruses’ genomic RNA, and then examined their persistence. Specifically, we measured first-order decay kinetics of an exogenous respiratory virus, SARS-CoV-2 RNA; an endogenous enteric virus, human norovirus (HuNoV) RNA; and two endogenous human fecal indicator viruses, pepper mild mottle virus (PMMoV) and tomato brown rugose fruit virus (ToBRFV) RNA in fecal sludge at three relevant temperature conditions (4 °C, 15 °C, and 30 °C). Collectively, these objectives contribute vital insights into fecal sludge sampling protocols in non-sewered settings and shed light on virus RNA persistence across diverse waste matrices. With the results of this study, we aim to advance understanding of monitoring infectious diseases in non-sewered settings.

## Methods

### Experiment overview and setup

Untreated fecal sludge was sampled from collection trucks at the inlet of a mid-sized fecal sludge treatment plant in Dakar, Senegal (Supplemental methods M1) and shipped to Stanford University, California. Each fecal sludge sample was aliquoted into duplicate 50-mL conical tubes. Tube racks were placed into three temperature-controlled rooms (4 °C, 15 °C, and 30 °C), and covered in aluminum foil to prevent photodegradation. The total experiment duration was 70 days, with samples taken throughout and analyzed for RNA of four target viruses (Figure S1).

### Site selection

In collaboration with Delvic Sanitation Initiatives, we identified six potential sampling sites in Senegal, considering operational areas, ease of sampling, and proximity to shipping services (Figure S2). Subsequently, we ranked and narrowed down these candidate regions based on COVID-19 prevalence in October 2020, ultimately choosing the city of Dakar. From the four FSTPs in Dakar, we selected the one with the largest “sewershed” population, serving approximately 600,000 people (Table S1). Cambérène, our chosen FSTP, receives 600 m^3^ of fecal sludge daily from emptying trucks servicing on-site sanitation systems and contains two parallel settling thickening tanks and 10 unpacked drying beds. The facility removes solids from the sludge before pumping the remaining liquid to a neighboring activated sludge wastewater treatment plant for further treatment (Figure S3).

### Target selection and screening

Candidate viruses were chosen through a comprehensive evaluation, considering factors such as relevance (e.g., COVID-19), regional importance, disease burden, knowledge gaps, logistical feasibility, and novelty of the work (Table S2). SARS-CoV-2, specifically the N1 and N2 regions of its genome, was selected due to its relevance to the COVID-19 pandemic. In 2017, diarrheal diseases, caused by enteric bacterial and viral pathogens, resulted in 1.6 million deaths globally, with children being the most affected (Dattani *et al*., 2023). Diarrheal diseases also rank among the top 5 causes of death in Senegal (*Senegal - CDC - Center for Global Health*, 2019). This prevalence motivated our selection of HuNoV, given its implications in a context where diarrheal diseases pose a considerable threat to public health.

Pepper mild mottle virus (PMMoV), a commonly used microbial source tracker (MST) found ubiquitously in feces, was included due to its use as a normalization basis for other WBE targets (D’Aoust *et al*., 2021; S. McClary-Gutierrez *et al*., 2021; Wu *et al*., 2022) and as an indirect measurement of the fecal strength of wastewater (Rosario *et al*., 2009; Symonds, Rosario and Breitbart, 2019). Although studies exist on PMMoV persistence in wastewater influent and solids (Roldan-Hernandez *et al*., 2022; Burnet *et al*., 2023; Li *et al*., 2023), there is novelty in studying PMMoV in on-site sanitation systems. Lastly, tomato brown rugose fruit virus (ToBRFV) was selected because it is an emerging microbial source tracker like PMMoV (Natarajan *et al*., 2023). ToBRFV is an RNA virus that infects tomatoes and peppers. Reports indicate its detection in crops in over 35 countries (Zhang *et al*., 2022), as well as its specificity and abundance in human feces (Rothman and Whiteson, 2022; Natarajan *et al*., 2023). This study marks the first detection of ToBRFV in Senegal and the first study globally to document the decay of ToBRFV RNA in any wastewater matrix based on a systematic literature search as of October 2023 (Supplemental methods M2 and M3).

Preliminary fecal sludge samples were sent to Stanford University for characterization in February and March 2022. Samples were screened for the presence of SARS-CoV-2, HuNoV, PMMoV, and ToBRFV virus RNA (assay information in Table S3). If a target was detected in the sacrificial screening sample, decay experiments were performed with the endogenous virus in the received fecal sludge. If no target was detected, exogenous virus was spiked before decay experiments. All three shipments (Table S4) of fecal sludge were negative for endogenous SARS-CoV-2 RNA during screening, which was expected considering the relatively low burden of disease in Dakar in the months preceding shipments (*Ministère de la santé, 2022*). All samples yielded detectable levels of PMMoV, ToBRFV and HuNoV RNA. Decay experiments proceeded with exogenous SARS-CoV-2 in the samples and the endogenous viruses (HuNoV, PMMoV and ToBRFV) found in the fecal sludge.

### Sample collection and processing

Grab samples were collected on October 3rd, 2022 from a vacuum truck during discharge of fecal sludge to Cambérène FSTP. Samples were manually composited; 3 L of samples were taken at the beginning, 5 L of sample in the middle, and 3 L of sample at the end of the discharge process. The sampling strategy was coordinated with vacuum truck operators and the samples were mixed vigorously with a stirring rod or ladle and collected to minimize settling (Figure S4). After collection in plastic bottles, sludge samples were enclosed in sealed plastic bags, placed in a styrofoam cooler, and shipped to Stanford University within 24 hours.

Upon arrival at Stanford (within 6 days), the samples were analyzed for temperature (20 °C), pH using a pH meter(Mettler Toledo, Columbus, -OH), and total solids using standard methods (U.S. Environmental Protection Agency, 2001); they were then stored at at 4 °C while the remaining experimental setup was completed for up to 12 hours. Samples were then mixed and aliquoted into duplicate 50 mL sterile conical tubes for each time point (Corning, Glendale, AZ); each conical tube was spiked with heat-inactivated SARS-CoV-2 (Integrated DNA technologies, Coralville, Iowa). First, a suspension of the SARS-CoV-2 stock was made by making a mix of 1 μL of stock per 1 mL of 1x tris-EDTA (TE) buffer. Then, 1 mL of the working stock was added to each aliquot for a resulting spike of (2.08 × 10^6^ gene copies per mL). Duplicate aliquots for each time point were subsequently incubated in three temperature-controlled rooms (4, 15, and 30 °C) to assess the effect of temperature on SARS-CoV-2, HuNoV, PMMoV, and ToBRFV RNA. The samples were covered in foil to prevent potential photodegradation. Both replicate aliquots were processed on day 0 (the day they were prepared), and on days 2,4,6,8,10,12,18,40,55, and 70 were immediately centrifuged upon collection for sample dewatering.

### RNA extraction and RT-ddPCR

We dewatered fecal sludge samples using a modified existing protocol and extracted them upon day of collection (Topol *et al*., 2021). After centrifugation at 24000 x g for 30 min at 4 °C, the entire supernatant was decanted, taking care not to disturb the pellet, and all solids were retained. 0.22 g of the dewatered sludge pellet were resuspended in a 2.9 mL solution of DNA/RNA shield spiked with 1.5 μL of rehydrated bovine coronavirus (BCoV) vaccine (500,000 copies/mL in the solution) as an extraction control. The final concentration of dewatered solids resuspended in the BCoV-spiked DNA/RNA shield was approximately 75 mg/mL. Homogenization with four grinding balls (OPS Diagnostics, Lebanon, NJ) at 1,000 rpm for two minutes (MP Biomedicals, Santa Ana, California) was followed by brief centrifugation at 4000 x g for five minutes. RNA extraction, conducted the same day, used Qiagen Allprep Powerviral Kits (Qiagen, Germantown, MD). Each sample was processed in duplicate from concentration through quantification. One positive and two negative extraction controls per sampling timepoint were included. RNA recovery efficiency was determined by comparing measured BCoV concentrations to stock concentrations, which also served as our positive extraction control.

Extracted RNA, stored at −80 °C (69−189 days), was quantified via reverse transcription droplet digital PCR (RT-ddPCR) using Bio-Rad (Hercules, CA) droplet digital PCR kits (catalog no. 12013743). Reaction mix concentrations and thermal cycling conditions can be found in supplemental methods M4. Each ddPCR plate included one positive control and three no-template (negative) controls. SARS-CoV-2 positive controls were patient-derived RNA. The positive control for BCoV was RNA extracted from reconstituted BCoV vaccines. Positive controls for PMMoV, HuNoV and ToBRFV consisted of synthetic gene fragments. ToBRFV’s positive control was cloned in a plasmid vector (IDT); the rest were purchased directly as gBlock gene fragments (IDT). All negative controls were molecular grade water (Corning, Corning, NY). The N1 and N2 duplex assay and HuNoV and ToBRFV singleplex assays were conducted in triplicate wells, and the BCoV/PMMoV duplex assay was conducted in duplicate wells; replicate wells were merged for analysis. QuantaSoft and Quantasoft Analysis Pro (Bio-Rad) were used for ddPCR data analysis, where clusters were manually assigned to positive and negative controls per manufacturer instructions (Bio-Rad). Formation of at least 10,000 droplets was required, and a sample was considered positive for a target if three or more droplets were positive in the merged wells.

### Data analysis

Data were exported as CSV files from ddPCR Quantasoft software and dimensional analysis to convert concentrations per well to gene copies per gram of fecal sludge solids was conducted in Microsoft Excel (version 16.71). The remainder of data processing, plotting, and model fitting analyses were performed in R (version 4.1.2) using Rstudio (version 2022.07.1) (Supplemental methods M5). For ddPCR data, the error bars are reported as standard deviations from total error associated with technical replicates (merged wells). Experimental replicate concentrations were averaged prior to plotting and model fitting, and errors were propagated for upper and lower error bars separately (Supplemental methods M6).

The decay of all RNA targets in fecal sludge was analyzed using a first-order decay model (Equation 1). The decay rate constant, *k,* was calculated by linearizing and determining the slope (Equation 2). The time required to achieve a 90% (one log) reduction (T_90_) was calculated using Equation 3.

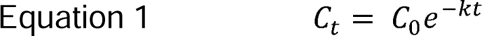

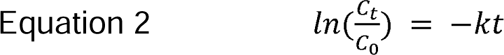

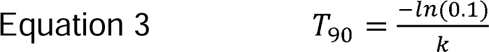

In these equations *C_t_*is the concentration of virus RNA at time *t*, *C_0_* is the starting concentration at time zero,and *k* is the first order decay rate constant.

## Results and discussion

### Quality assurance and quality control

We followed the Environmental Microbiology Minimum Information (EMMI) Guidelines (Borchardt *et al*., 2021) as closely as possible (Supplemental methods M7). Extraction and ddPCR positive and negative controls were positive and negative, respectively, for all viral RNA targets. BCoV recovery averaged 29%, and the limit of detection was determined by identifying samples with three positive droplets. Samples meeting this threshold were considered positive.

Graphical analysis of viral RNA target concentration assessed the impact of template dilution (both undiluted and 10x dilution were performed). RNA extracts were run at a tenfold dilution for all targets, with undiluted plates selected for SARS-CoV-2 targets due to samples falling below the limit of detection at a dilution factor of 10. Duplicate samples were averaged for plotting, except when one data point was missing. Three sample replicates were excluded from analyses due to lab processing loss. In such cases, the concentration value of the viable replicate was used.

### Overall detection of targets

Endogenous viruses PMMoV and ToBRFV are non-infectious to humans but their RNA is abundant in human feces and found at similar orders of magnitude (6.6 x 10^7^ and 7.67 x 10^7^ gc/g in initial samples, respectively, Figure 1A). They were found at concentrations almost 1.5 orders of magnitude higher than HuNoV (2.9 x 10^6^ gc/g in initial samples). This difference in abundance was expected because while all feces may contain fecal markers, not all individuals are infected with HuNoV. Exogenous (spiked) SARS-CoV-2 N1 and N2 genes were consistently detected at an average concentration of 3.51 x 10^4^ gc/g in initial samples. All targets remained detectable throughout the 70-day experiment (Figure 1B).

**Figure 1.**
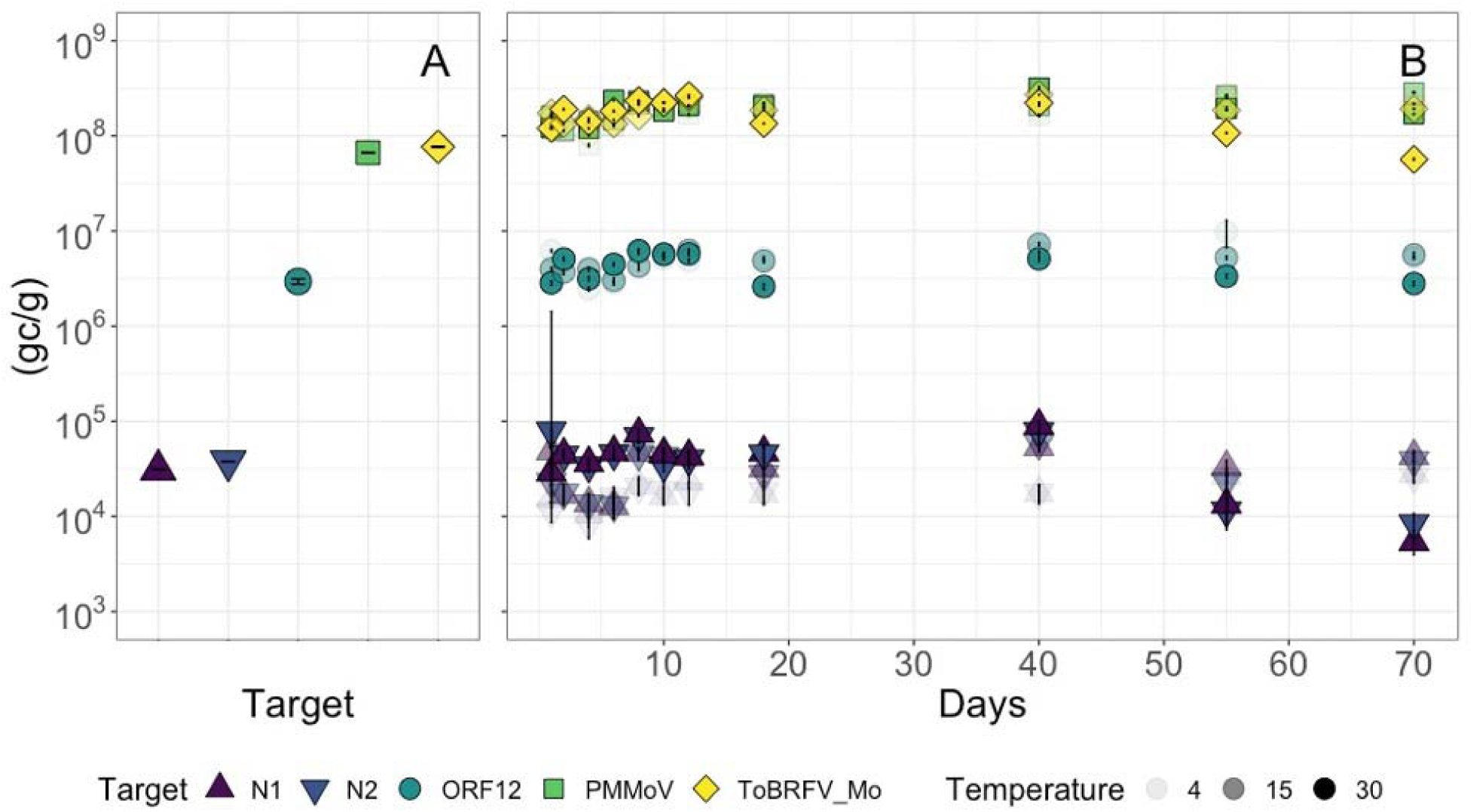
Panel A (left) illustrates the concentrations of all target viruses at 4, 15, and 30 °C in the initial sample, while panel B (right) depicts concentrations throughout the 70-day experiment reported at gene copies per gram of dry fecal sludge solids. In the legend, SARS-CoV-2 targets (N1 and N2) are denoted by triangle points, HuNoV (ORF12) by circles, and MST viruses (PMMoV and ToBRFV) by squares. The opacity of the data points in Panel B increases with temperature. Error bars depict standard deviations among experimental duplicates (aliquots) and technical replicates (wells) for each target.

### Persistence study

Despite the extended experiment duration, limited decay (<1 log_10_ reduction) was observed for all RNA targets across all temperature conditions. Table 1 provides details on the first-order decay rate constants (hereafter referred to as “*k* values”) and the time required to achieve a 90% reduction (T_90_) in concentration for SARS-CoV-2 (N1 and N2 genes), HuNoV, PMMoV, and ToBRFV. Positive *k* values were exclusively observed at 30° C (p < 0.05), ranging from 0.010 to 0.023 day^-1^. However, at 4 and 15 °C, *k* was negative (indicating increasing concentration over time) for all RNA targets, although often not significantly different from zero (Table 1). Additional linearized data for model fits can be found in supplemental table S6.

**Table 1.** First-order decay rate constants k for all targets and all temperatures in fecal sludge. Negative k values indicate there was a positive slope (indicating C > C_o_); T_90_ values were not calculated for positive slopes. Asterisk (*) indicates k values significantly different from zero (p < 0.05).

| Target | Temperature (°C) | *k* (days<sup>-1</sup>) | *p* value | *T*<sub>90</sub> (days) | *T*<sub>90</sub> (months) |
| --- | --- | --- | --- | --- | --- |
| N1 | 4 | -0.01 | 0.02* |  |  |
|  | 15 | -0.01 | 0.27 |  |  |
|  | 30 | 0.023 | 0.02* | 100 | 3.3 |
| N2 | 4 | -0.01 | 0.02* |  |  |
|  | 15 | -0.01 | 0.24 |  |  |
|  | 30 | 0.023 | 0.01* | 100 | 3.3 |
| HuNoV | 4 | -0.008 | 0.14 |  |  |
|  | 15 | -0.01 | 0.07 |  |  |
|  | 30 | 0.01 | 0.32 | 500 | 16.7 |
| PMMoV | 4 | -0.01 | 0.02* |  |  |
|  | 15 | -0.01 | 0.004* |  |  |
|  | 30 | -0.003 | 0.50 |  |  |
| ToBRFV | 4 | -0.007 | 0.09 |  |  |
|  | 15 | -0.004 | 0.23 |  |  |
|  | 30 | 0.012 | 0.03* | 187 | 6.2 |

### N1/N2 targets

Exogenous SARS-CoV-2 RNA exhibited high persistence, with statistically significant decay (k=0.023 and p<0.05 for N1 and N2) only observed at 30 °C, resulting in a T_90_ value of 3.3 months (Table 1). At 15 °C, minimal to negligible decay was observed. In fact, the RNA concentrations remained stable (*k* = -0.01 day^-1^, p > 0.05). At 4 °C there was a slight positive slope (Figure 2A and B) that was significantly different from 0 (*k* = -0.01 day^-1^ and p < 0.05), perhaps due to the small sample size (12 time points) of the study.

**Figure 2.**
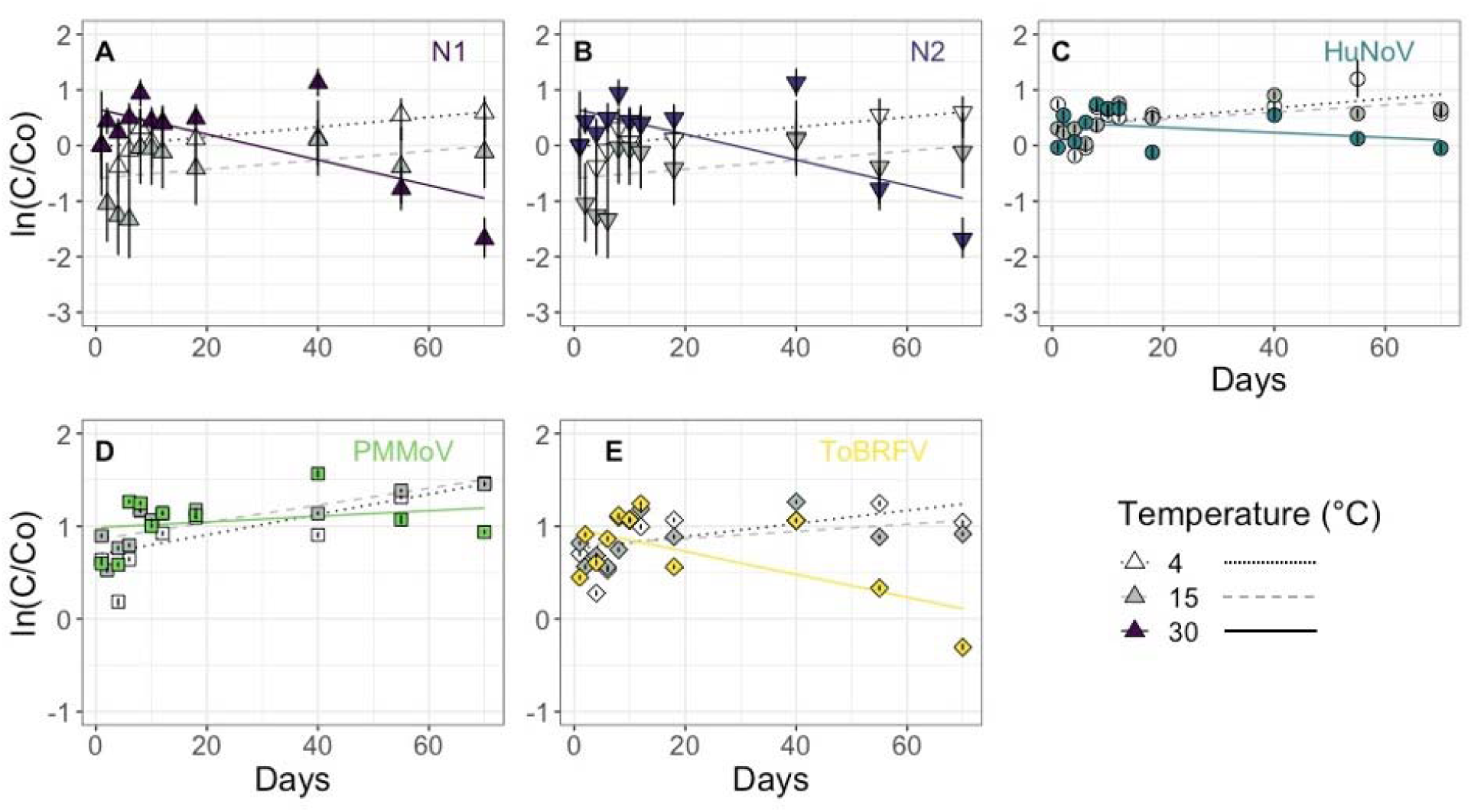
First-order decay curves of SARS-CoV-2 N1 (A), N2 (B), HuNoV (C), PMMoV (D) and ToBRFV (E) RNA in fecal sludge. Temperature is indicated by open (4 °C), gray (15 °C), and solid color (30 °C) symbols with dotted, dashed, and solid lines, respectively. Colors and shapes indicate target type consistent with Figure 1. Error bars depict standard deviations among experimental duplicates (aliquots) and technical replicates (wells) for each target.

Our findings share several parallels with existing observations. So far, only one other persistence study, conducted using river water and seawater at 4 and 20° C, also observed negligible decay of SARS-CoV-2 RNA (Sala-Comorera *et al*., 2021). Persistence studies in wastewater influent report higher *k* values with substantially faster decay at higher temperatures, ranging from *k* = 0.05 day^-1^ at 4 °C to as high as 4.32 day^-1^ at 20°C (Weidhaas *et al*., 2021; Burnet *et al*., 2023). However, a study by Roldan-Hernandez (2022), focusing on persistence in primary settled solids from a centralized system in the US, observed a slower decay with *k* values approaching levels comparable but higher than those in the present study (*k* = 0.024 day^-1^ at 4°C, and *k* = 0.06 day^-1^ at 37°C). This convergence may be attributed to the higher solids content shared by primary settled solids and fecal sludge (Table S5), underscoring the potential influence of matrix composition on decay dynamics.

### HuNoV target

No significant decay (p > 0.05) was observed for HuNoV RNA at any of the three temperature conditions (Figure 2C). Few studies investigate the persistence of HuNoV RNA, and those that do have focused on inactivation or detection in environmental waters—a justified emphasis given its contribution to recreational waterborne illness (Viau, Lee and Boehm, 2011; Boehm, Soller and Shanks, 2015; Wade *et al*., 2018). Nevertheless, there is a lack of attention to persistence in wastewaters, including fecal sludge. Our results align with other studies despite differences in the water matrix. One recent study conducted in river water documented a range of *k* values for HuNoV RNA, spanning from no significant decay to *k* = 0.12 day^-1^ (Kennedy *et al*., 2023).

These persistence results are impactful for water, sanitation, and hygiene (WASH) initiatives because the provision of universally safe sanitation directly influences disease burden (World Health Organization and United Nations Children’s Fund (UNICEF), 2017). For example, one study conducted in Dhaka, Bangladesh revealed a 14% lower prevalence of diarrheal disease in communities with access to safely managed sanitation compared to those without (Akter *et al*., 2022). At present, the high persistence of HuNoV RNA in fecal sludge suggests a feasible opportunity to collect public health data from on-site sanitation contexts. Although HuNoV RNA remains detectable via molecular methods for longer than viable norovirus (Shaffer *et al*., 2022), future studies should elucidate exposure risks to sanitation workers and the potential impacts on broader public health initiatives.

### PMMoV and ToBRFV targets

Based on our literature review, we hypothesized minimal decay of PMMoV RNA; indeed, there was no significant decay even at 30 °C (*k* = -0.003 day^-1^, p > 0.05). Roldan-Hernandez *et al*. (2022) found high persistence of endogenous PMMoV RNA in primary settled solids, with decay rates varying between 0.01 and 0.09 day^-1^ (Table 1). At 4 °C and 15 °C, RNA concentrations in final samples after 70 days were slightly but statistically significantly higher than the initial concentrations (Figure 2D). This observed variation is likely attributable to measurement artifacts that reflect inherent variability across all sample processing stages. Burnet et al. (2023) also observed increased PMMoV RNA concentrations at 4, 12, and 20 °C after 27 days. Other hypotheses for these observations include potential changes in the composition (biotic or abiotic) of the sample matrix during incubation, which might enhance the availability of RNA for extraction and result in variable decay rate constants (Burnet *et al*., 2023; Kennedy *et al*., 2023). Observing similar persistence as previous reports demonstrates PMMoV’s utility as both a microbial source tracker and a fecal strength control in various contexts.

ToBRFV RNA was overall highly persistent over 70 days at the lower temperatures of 4 and 15 °C. At 30 °C (Figure 2E), slight but significant decay was observed (*k* = 0.012 day^-1^, T_90_ = 6.2 months). Although the literature lacks comparative *k* values, our results describing faster decay at higher temperatures are consistent with general trends for other targets (e.g., PMMoV). To date, ToBRFV has been suggested as an MST tool due to its specificity to humans and abundance in feces. In the broader context of WBE, particularly for on-site systems, PMMoV and ToBRFV serve as highly resilient fecal indicator viruses.

### Implications

The decay rate constants, on the whole, are quite modest; *k* values for all targets in our study consistently show lower values than those reported for sewered wastewater influent and wastewater solids. However, they are consistent with findings indicating that virus RNA exhibits minimal decay in wastewater solids (Roldan-Hernandez *et al*., 2022; Burnet *et al*., 2023). The limited decay observed implies that the targets remain detectable for an extended period. This finding is encouraging for using FSTPs as a centralized point for sampling and identifying viral targets and underscores the importance of understanding average time between collections in non-sewered settings (Delaire *et al*., 2021).

In some studies, nonlinear decay models, such as JM2 (Juneja, Huang and Marks, 2006), have been proposed for pathogens and fecal indicators in wastewater, manure, and biosolids, as observed by Mitchell (2015). This model accommodates shouldering (an initial delay in decay) and tailing (decay slowing with time). Considering these features, our findings of exceptionally low decay rates for endogenous viral RNA targets might be explained by capturing the tail end of decay models seen in other studies, making our observed decay curve appear relatively flat. Capturing the tail end is logical considering fecal sludge from emptying trucks exhibits a mixed age profile due to variations in filling durations and emptying rates among on-site sanitation systems (Conaway *et al*., 2023). However, a prior study in Malawi found that sampling depth in pit latrines (indirect measure of sludge age) did not strongly impact the detection of enteric pathogens using molecular methods (Capone *et al*., 2021), suggesting consistent detection even in aged fecal sludge.

### Limitations

Several limitations should be considered to contextualize the results of this study. First, this study is limited to RNA persistence, not infectivity. Consequently, the findings do not necessarily imply potential risks to sanitation workers, despite the significance of this concern. It is also possible that by sampling at an FSTP where fecal sludge has been pumped out of containment, we may have exposed the sample to aerobic conditions and thus disrupted ongoing anaerobic processes. Additionally, the 4 to 5 days of sample shipment time may lead to differences between our initial concentrations of endogenous viral RNA and the concentrations in feces deposited into the pit latrines from which the sludge was collected. This lag allows for the possibility of biological or chemical processes in the initial days before analysis, potentially resulting in decay not captured in our study. Future research can estimate decay during transport by introducing a known concentration of a tracer virus at the time of sampling and measuring its concentration immediately upon sample receipt after shipment.

Because our study was conducted using a composited sample from one fecal sludge collection truck, future work should also look at inter-shipment variability and seasonal variability of viral targets in fecal sludge. This approach mirrors the methodology employed in studies within sewered settings, where intraday variability in wastewater influent is commonly examined (Ahmed *et al*., 2021; Bivins *et al*., 2021; Mendoza Grijalva *et al*., 2022). Further decay experiments are necessary for fecal sludges at temperatures exceeding 30°C, which are typical in regions like sub-Saharan Africa and India. Additionally, experiments conducted at room temperature (20°C) can address potential limitations in accessing temperature-controlled facilities or incubators, which may vary both in the field and among different laboratories. While our study indicates that refrigerating samples may not be necessary for accurate detection and quantification of viruses, it is advisable to verify our findings. Valuable insights into workflow optimization can be obtained through such investigations.

## Conclusions

This study introduces an original approach to WBE in a non-sewered area in Dakar, Senegal. We collected fecal sludge samples from a centralized facility that receives and consolidates fecal sludges from nearby on-site sanitation systems. Our analysis identified the presence of endogenous HuNoV, PMMoV, and ToBRFV RNA. Exogenous SARS-CoV-2 N1 and N2 RNA exhibited modest decay at 30 °C (k = 0.023 day^-1^, p < 0.05), corresponding to a T_90_ value of 3.3 months. Endogenous ToBRFV RNA also showed modest decay at 30 °C (k = 0.012 day-1, p < 0.05), with a T_90_ value of 6.2 months. No significant decay was observed for HuNoV or PMMoV RNA at any temperature condition (4, 15, and 30 °C) over the 70-day period. Our study presents the first detection of ToBRFV as a novel microbial source tracker in Senegal wastewaters, and the first persistence study of its nucleic acids in any water matrix. By providing novel first-order decay rate constants, this study begins to address a significant knowledge gap concerning the stability of viral RNA in fecal sludges.

Next steps for assessing the utility of sampling from FSTPs for community health surveillance include examining correlations between clinical data and fecal sludge data from FSTPs, as has been done with environmental surveillance studies (Rogawski McQuade *et al*., 2023). Utilizing FSTPs could streamline site selection for environmental surveillance studies because they are already centralized and contained, which could facilitate data interpretation. Sampling directly from on-site sanitation systems offers more control over obtaining fresh sludge due to sampling depth (Capone *et al*., 2021; Li *et al*., 2023), while opting for older sludge at an FSTP may provide a more representative sample of a wider community. Overall, results from our study suggest the feasibility of utilizing FSTPs for WBE sampling and highlight their potential as a valuable complement to current surveillance of wastewater and environmental waters. On-site sanitation technologies constitute up to 72% of sanitation system usage in much of the global south (Greene et al., 2021); our findings encourage researchers to leverage established fecal sludge management infrastructure to develop community health surveillance programs.

## Data Availability

All data produced are available in the manuscript or available upon request.

## Acknowledgments

Financial support for this study was generously provided by the Stanford Diversifying Academia, Recruiting Excellence (DARE) Fellowship awarded to LMG, as well as grants from the Stanford School of Engineering, the Dreyfus Foundation, the Stanford Center for Innovation in Global Health (CIGH), and the Stanford Department of Chemical Engineering. We would also like to acknowledge the team at Delvic Sanitation Initiatives for sample collection and Aravind Natarajan for his technical assistance with the ToBRFV assay. Graphical abstract was created using BioRender.

## Supplementary materials

The supplementary materials document contains details about experimental design, literature review process, and data processing.

