## Supplemental Information for "Persistence of respiratory, enteric, and fecal indicator viruses in fecal sludge from on-site sanitation in Dakar, Senegal"

**Description of Supplementary Materials**

Number of pages: 22

Number of methods: 6

Number of figures: 5

Number of tables: 5

Table of contents

| **Supplemental methods** | Page |
| --- | --- |
| M1. Sample collection | 4 |
| M2. Systematic literature review process for virus decay | 4 |
| M3. Systematic literature review process for non-sewered WBE | 5 |
| M4. Further information for ddPCR | 6 |
| M5. Dimensional analysis for converting ddPCR instrument output to desired units | 6 |
| M6. Propagation of error | 8 |
| M7. EMMI Guidelines | 9 |
| **Supplemental Figures** |  |
| Figure S1. Experimental workflow graphic | 10 |
| Figure S2. Candidate regions in Dakar for FSTP site selection | 11 |
| Figure S3. Cambérène fecal sludge treatment plant (FSTP) and wastewater treatment plant (WWTP) process flow diagram | 12 |
| Figure S4. Sampling from pumping trucks | 13 |
| **Supplemental Tables** |  |
| Table S1. Senegal FSTPs “sewer”shed population | 14 |
| Table S2. Summary or viral RNA targets selected for this study and their justification | 15 |
| Table S3. Table S2. RT-ddPCR assays | 16 |
| Table S4. Table S3. Fecal sludge shipment schedule | 18 |
| Table S5. Solids content of several wastewater matrices | 19 |
| Table S6. Linearized model fits for virus RNA decay | 20 |

### Supplemental Methods

#### M1. Sample collection and shipping

Grab samples were collected on October 3rd, 2022 from a vacuum truck during discharge of fecal sludge to Cambérène FSTP. Samples were manually composited; three liters (3 L) of samples were taken at the beginning, 5 L of sample in the middle and 3 L of sample at the end of the discharge process. The sampling strategy was coordinated with the vacuum truck operators and the samples were mixed vigorously with a stirring rod or ladle and collected to minimize settling in the bucket (Figure S5). Samples were placed in coolers with wet ice and mailed to Stanford. Typical transit times ranged from 4-5 days from day of collection. Upon arrival, samples were stored at 4 ^o^C until processed.

#### M2. Systematic literature review process for virus decay

On October 4th 2023, a systematic literature review was conducted to find persistence studies of SARS-CoV-2, HuNoV, PMMoV, and ToBRFV in wastewater. The following Boolean search terms/operators were used:(Decay OR Persistence OR Stability) AND (Wastewater OR Sewage OR Septage OR Sewer* OR “fecal sludge” OR “faecal sludge” OR Influent OR Solids) AND (Virus* OR SARS-CoV-2 OR Coronavirus* OR Norovirus* OR PMMoV OR Pepper Mild Mottle Virus OR ToBRFV OR “Tomato brown rugose fruit virus”). The same search terms were applied in three databases (PubMed, Scopus, and Web of Science) for the generation of papers. All English results published were taken into consideration. Covidence was employed to import references, exclude duplicates, and facilitate screening processes with filtering features. In the criteria for inclusion, papers with primary experimental results and those addressing any of the target viruses outlined in the boolean search string were considered. Exclusion criteria involved the omission of papers solely investigating viability (culturable virus) without employing molecular methods. Additionally, papers in matrices deemed irrelevant, such as food, were excluded. Also excluded were literature reviews, along with studies focusing on "surrogate" viruses, those utilizing secondary data, and investigations into the effects of freezing and thawing on viruses at negative temperatures. Exclusive focus was placed on the predefined target viruses. 1361 studies were initially imported, and 303 duplicates were removed. 1058 studies were screened by title and abstract for relevance; 77 full-test studies were assessed for eligibility, and a total of 20 studies fit the inclusion criteria and reported *k* values. Those 20 studies were retained to use as source material.

#### M3. Systematic literature review process for non-sewered WBE

On October 4th 2023, a systematic literature review was conducted to identify and evaluate studies conducting WBE in non-sewered settings. The following boolean search terms/operators were used: (WBE OR “wastewater-based epidemiology” OR “wastewater surveillance”) AND (“non-sewered” OR “onsite sanitation” OR septic OR “pit latrine” OR rural OR LMIC). The same search terms were applied in three databases (PubMed, Scopus, and Web of Science) for the generation of papers. All English results published were taken into consideration. Covidence was employed to import references, exclude duplicates, and facilitate screening processes with filtering features. In the criteria for inclusion, papers presenting primary experimental findings, particularly those focusing on molecular investigations of pathogens in regions with either limited or high-income countries lacking centralized sanitation infrastructure were considered. Exclusion criteria involved the omission of studies examining environmental waters immediately downstream from centralized wastewater treatment facilities. Also excluded were studies from low-income countries *with* sewered or centralized wastewater treatment. 174 studies were initially imported, and 82 duplicates were removed. 92 studies were screened by title and abstract for relevance; 43 full-text studies were assessed for eligibility and a total of 8 studies fitting the inclusion criteria were retained to use as source material.

#### M4. Further information for ddPCR

#### ddRT-PCR was performed on 20 μl samples from a 22 μl reaction volume, prepared using 5.5 μl template, mixed with 5.5 μl of One-Step RT-ddPCR Advanced Kit for Probes (Bio-Rad 1863021), 2.2 μl of 200 U/μl Reverse Transcriptase, 1.1 μl of 300 mM dithiothreitol (DTT) and primers and probes mixtures at a final concentration of 900 nM and 250 nM respectively. Droplets were generated using an AutoDG (BioRad) using automated droplet generation oil for probes (BioRad; cat. no.1864110). Once droplets were generated, plates were sealed and placed onto a thermocycler within 30 minutes of generation.

#### N1/N2, HuNoV and ToBRFV assay plates were thermocycled as follows: 50°C for 60 minutes, 95°C for 10 minutes, 40 cycles of 94°C for 30 seconds then 55°C for 1 minute, followed by 98°C for 10 minutes, and 4°C for at least 30 minutes. PMMoV/BCoV plates were thermocycled as follows: 50°C for 60 minutes, 95°C for 10 minutes, 40 cycles of 94°C for 30 seconds then 56°C for 1 minute, followed by 98°C for 10 minutes, and 4°C for at least 30 minutes. Plates were moved to a QX200 droplet reader (BioRad) within 48 hours of thermocycling.

#### M5. Dimensional analysis for converting ddPCR instrument output to desired units

Concentrations per reaction were converted into gene copies per gram of dry fecal sludge solids (gc/g) using the following dimensional analysis:

Equation 1	$C_{merged} = C_{exp} x V_{merged})$

Equation 2	$C_{extract} = \frac{C_{merged}}{V_{rxn}} x D$

Equation 3	$C_{final} = \frac{C_{extract} x V_{eluted}}{M_{dry}}$

C_merged = RNA concentration of merged triplicate reaction (gc per well)

C_exp = Concentration exported from plate reader (gc/uL)

V_merged = Volume of merged 20 uL wells (60 uL if in triplicate, 40 uL if in duplicate)(uL)

C_extract = RNA concentration in template from extraction kits (gc/uL)

C_rxn = RNA concentration in merged reaction wells (gc/uL)

V_rxn = vol of RNA input per reaction (per well) (uL)

D = dilution factor. (e.g., 1 if undiluted template for reaction or 10 for a 1:10 dilution of RNA template)

C_extract = RNA extract concentration (gc/uL)

V_eluted = Total volume of RNA eluted from extraction kit (uL)

M_dry = Mass of dry fecal sludge solids (g)

C_final = final RNA concentration in mass of dry fecal sludge solids (gc/g)

#### M6. Propagation of error

Equation 1	$\delta Z = \frac{1}{2}\sqrt{\delta A^{2}+\delta B^{2}}$

Where $\delta Z$ represents the error associated with the final averaged value, $\delta A$ represents the error associated with one experimental replicate, and $\delta B$ represents the error associated with the second experimental replicate. The error associated with each experimental replicate (50-mL conical vial) arose from merging three replicate ddPCR wells (technical replicates). For creating the decay plot where concentrations at a certain point in time were divided by the concentration at time zero, error was propagated according to equation 2.

Equation 2	$\delta(\frac{a}{b}) = \frac{a}{b}\sqrt{{\frac{\delta a}{a}}^{2}+{\frac{\delta b}{b}}^{2}}$

Where *a* is the concentration of any given target at a particular time during the experiment, *b* is the concentration of that target at time T = 0, and $\delta$ denotes the error associated with each value, which includes the error from both duplicate experimental replicates and triplicate wells.

#### M7. Details requested in the EMMI Guidelines checklist

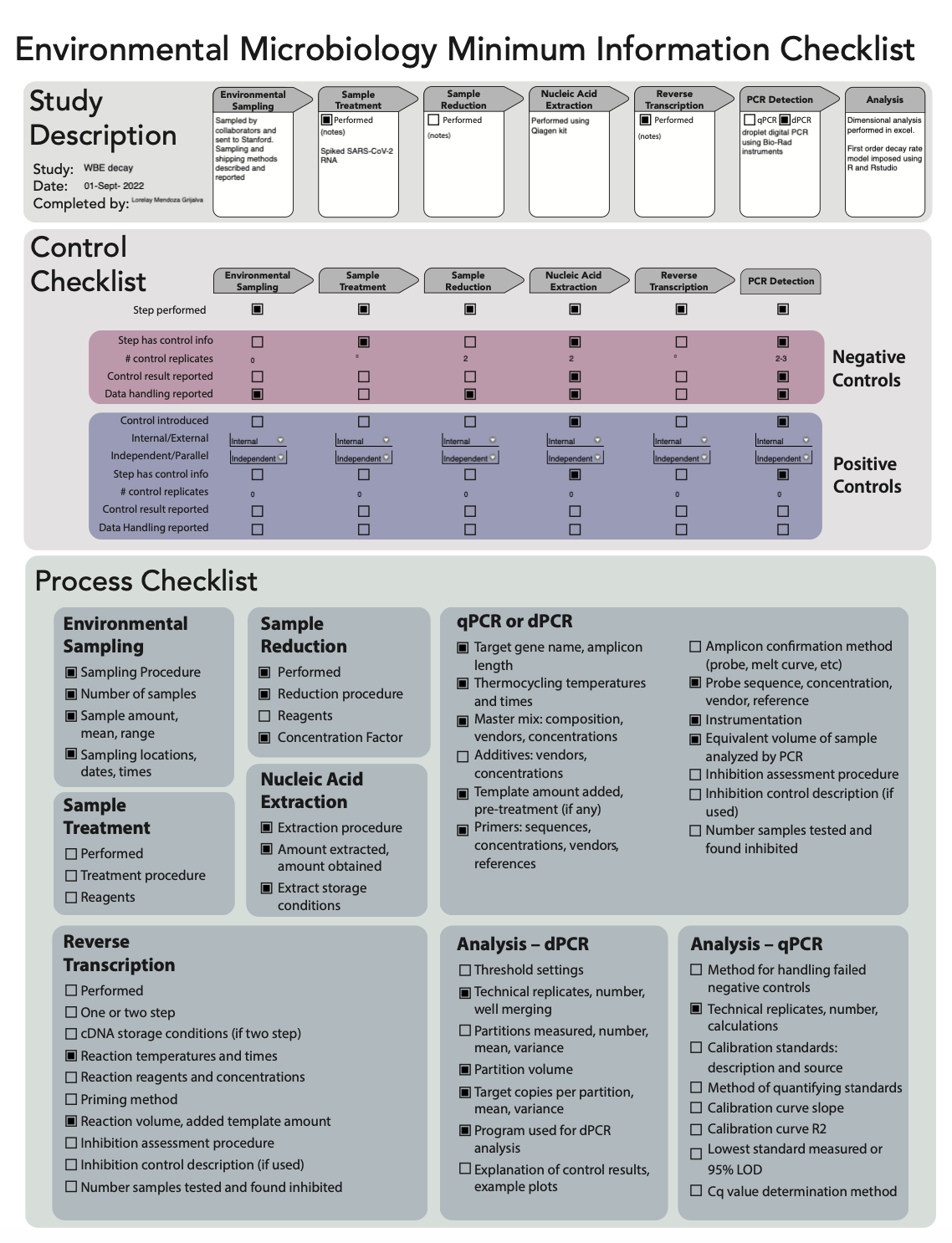

### Supplemental Figures

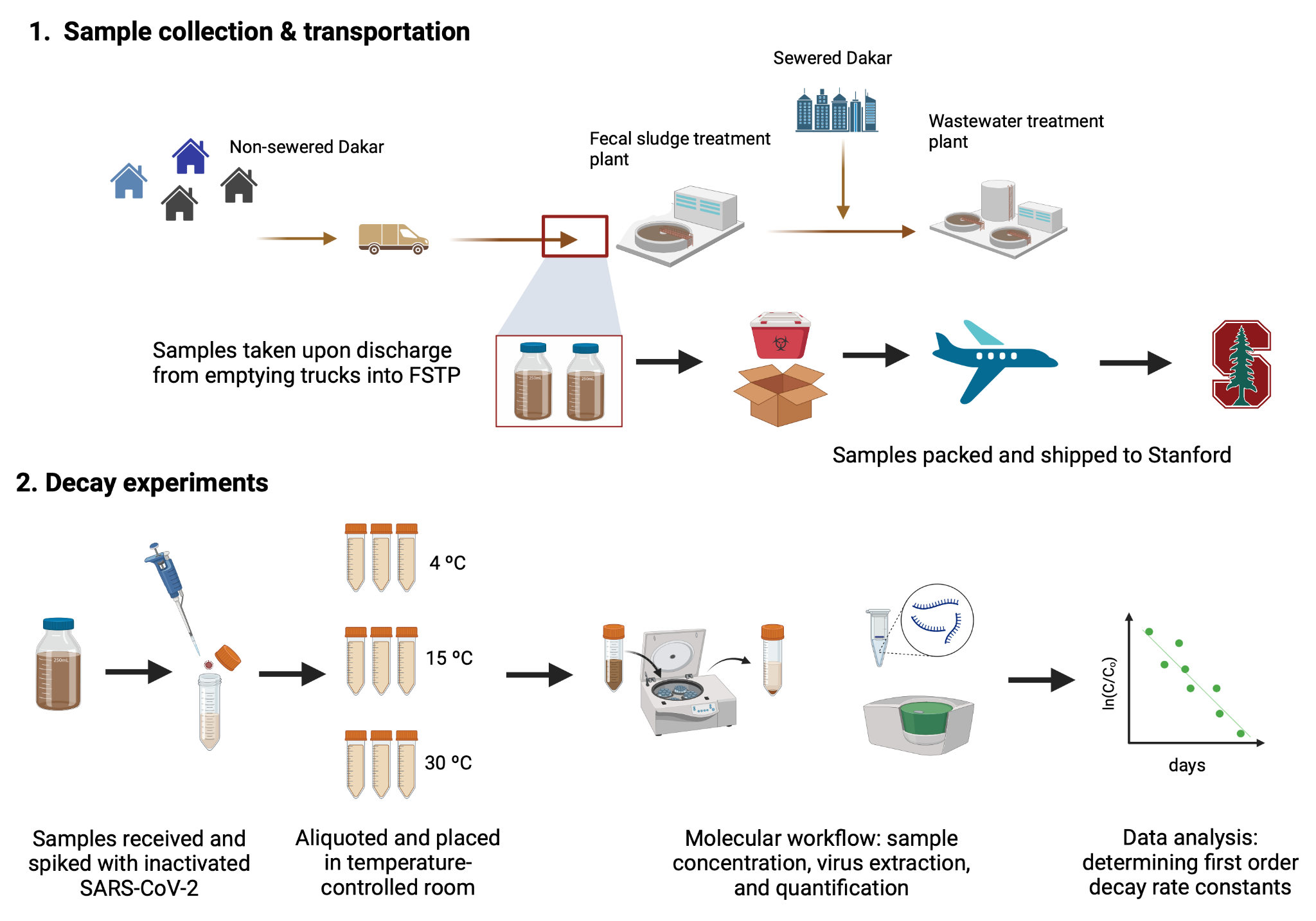

#### Figure S1. Experimental workflow graphic

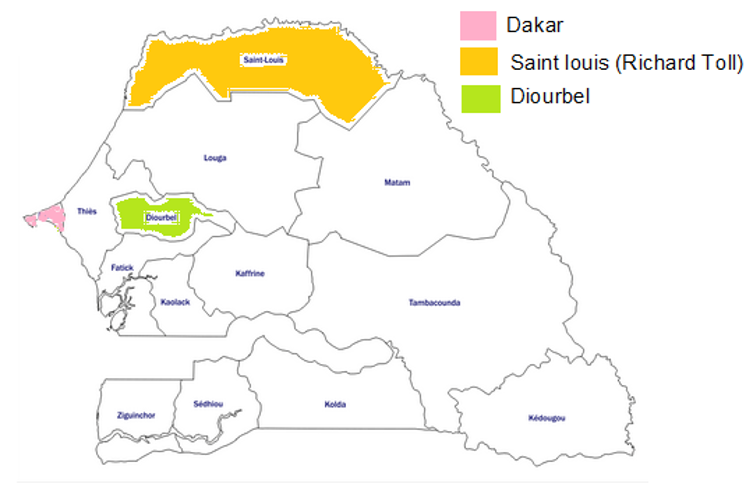

#### Figure S2. Candidate regions in Dakar for FSTP site selection

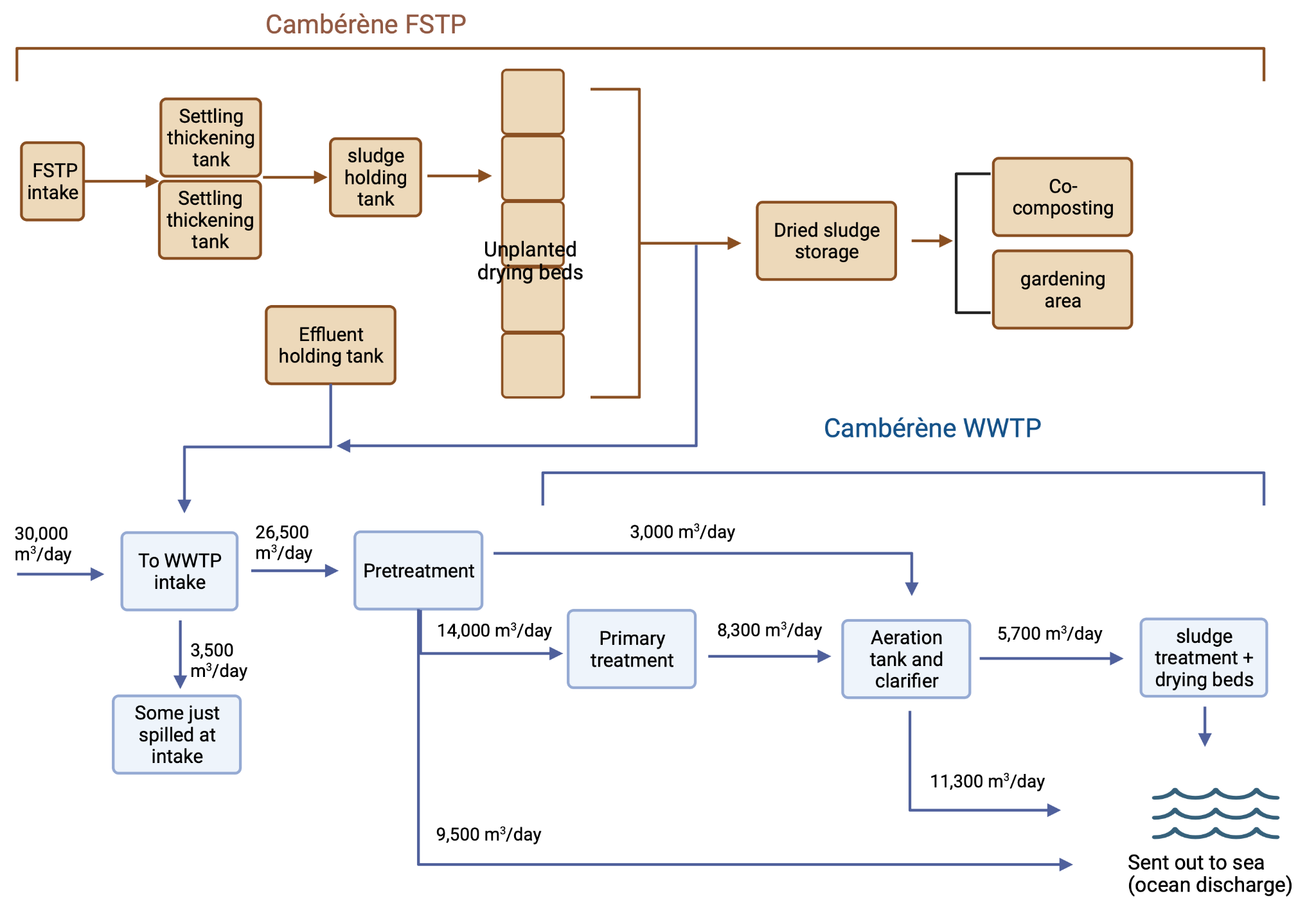

#### Figure S3. Cambérène fecal sludge treatment plant (FSTP) and wastewater treatment plant (WWTP) process flow diagram

The treatment consists of two parallel settling thickening tanks and 10 unplanted drying beds. The effluent from the settling thickening tanks flows to the effluent holding tank before being pumped to the neighboring wastewater treatment plant (activated sludge).

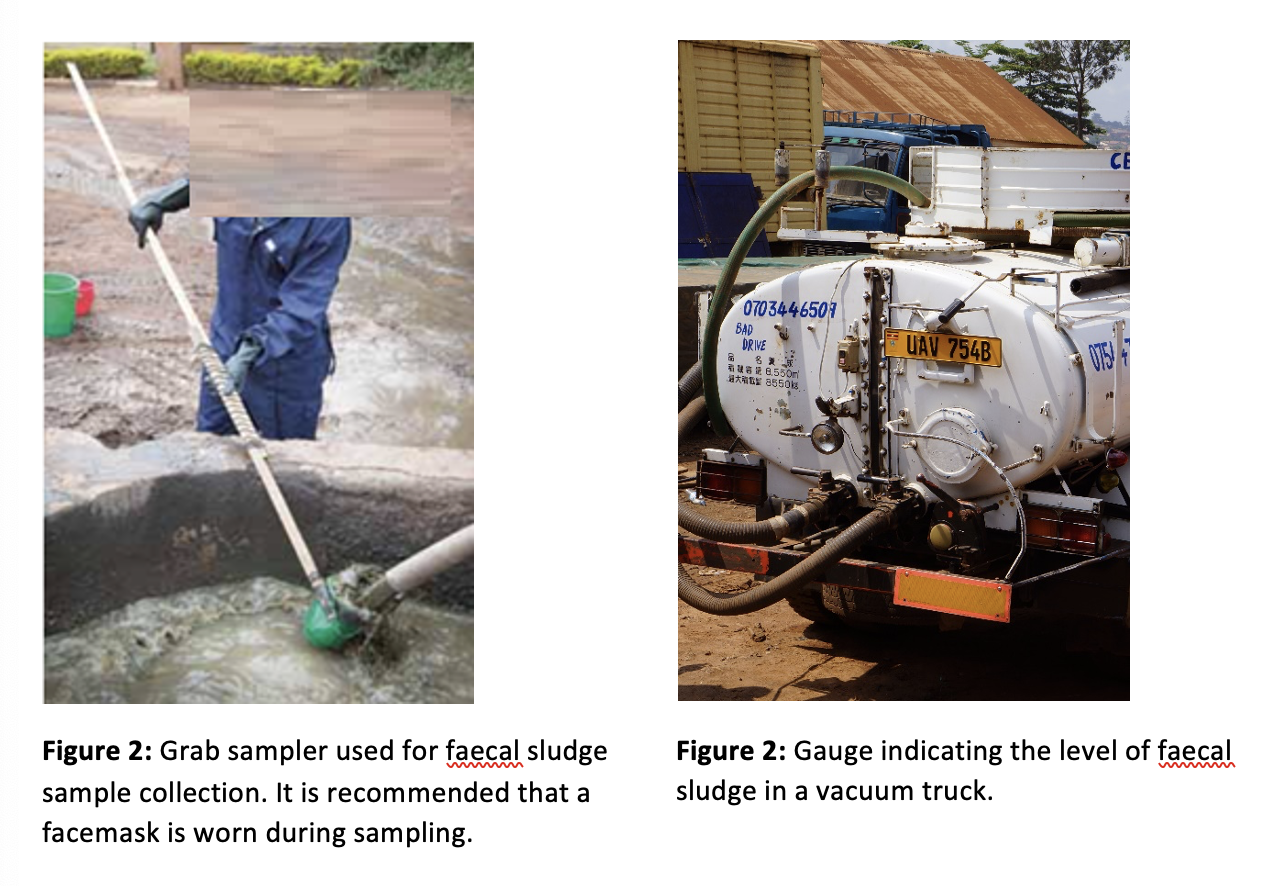

#### **Figure S4 :** Sampling from pumping trucks

### Supplemental Tables

#### Table S1. Senegal FSTPs “sewer”shed population

| **Region** | **City** | **Population** |
| --- | --- | --- |
| Dakar | Cambérène | 600,000 |
|  | Pikine (Niayes) | 500,000 |
|  | Tivaouane Peulh | 150,000 |
|  | Rufisque | 500,000 |
| Diourbel | Diourbel | 63,300 |
| Saint-Louis | Richard Toll | 13,000 |

#### Table S2. Summary or viral RNA targets selected for this study and their justification

| **Target** | **Infection type** | **Source** | **Justification** | **Assay** |
| --- | --- | --- | --- | --- |
| SARS-CoV-2 N1 | Respiratory | Exogenous  (heat inactivated) | Of interest due to COVID-19 pandemic | (Lu et al. 2020) |
| SARS-CoV-2 N2 | Respiratory | Exogenous  (heat inactivated) | Of interest due to COVID-19 pandemic | (Lu et al. 2020) |
| Human norovirus (HuNoV) | Enteric | Endogenous | Leading cause of diarrheal death in LMICs | (Loisy et al. 2005; Kennedy et al. 2023) |
| Pepper mild mottle virus (PMMoV) | Human fecal indicator virus | Endogenous | Established MST and WBE normalizer | (Haramoto et al. 2013) |
| Tomato brown rugose virus (ToBRFV) | Human fecal indicator virus | Endogenous | Novel MST | (Natarajan et al. 2023) |

#### Table S3. RT-ddPCR assays

| **Assay Target** | **Primer/probe sequence (5’-3’)** | | **Amplicon length (bp)** | **Reference** |
| --- | --- | --- | --- | --- |
| SARS-CoV-2 N1 | Forward | GACCCCAAAA TCAGCGAAA T | 72 | (Lu et al. 2020) |
|  | Reverse | TCTGGTTACTGCCAGTTGAATCTG |  |  |
|  | Probe | ACCCCGCATTACGTTTGGTGGACC (5' FAM/ZEN/3' IBFQ) |  |  |
| SARS-CoV-2 N2 | Forward | TTACAAACATTGGCCGCAAA | 67 | (Lu et al. 2020) |
|  | Reverse | GCG CGACA TTCCGAAGAA |  |  |
|  | Probe | ACAA TTTGCCCCCAGCGCTTCAG (5' HEX/ZEN/3' IBFQ) |  |  |
| Norovirus GII | Forward | ATGTTCAGRTGGATGAGRTTCTCWGA |  | (Kennedy et al. 2023; Loisy et al. 2005) |
|  | Reverse | TCGACGCCATCTTCATTCACA |  |  |
|  | Probe | AGCACGTGGGAGGGCGATCG (FAM/IBHQ) |  |  |
| BCoV | Forward | CTGGAAGTTGGTGGAGTT | 85 | (Decaro et al. 2008) |
|  | Reverse | ATTATCGGCCTAACATACATC |  |  |
|  | Probe | CCTTCATATCTATACACATCAAGTTGTT (5' FAM/ZEN/3' IBFQ) |  |  |
| PMMoV | Forward | GAGTGGTTTGACCTTAACGTTTGA | 68 | (Haramoto et al. 2013; Zhang et al. 2005) |
|  | Reverse | TTGTCGGTTGCAA TGCAAGT |  |  |
|  | Probe | CCTACCGAAGCAAATG (5' HEX/ZEN/3' IBFQ) |  |  |
| ToBRFV | F | TCA GTG TCT GTT TGG TCG ATA A |  | (Natarajan et al. 2023) |
|  | R | GGA ACG ACT TTG AAC TGA AAC C |  |  |
|  | P | AGA GCG GAC GAG GCA ACT CTT G  (FAM/ZEN/IBH Q) |  |  |

#### Table S4. Fecal sludge shipment schedule

| **Shipment** | **Date** | **Purpose** |
| --- | --- | --- |
| 1 | 2/4/2023 | Sacrificial sample for virus screening |
| 2 | 3/22/2023 | Sacrificial sample for virus screening |
| 3 | 10/10/2023 | Sample for experiments |

#### Table S5. Solids content of several wastewater matrices

| **Source** | **Matrix** | **Solids content** | **Solids type** |
| --- | --- | --- | --- |
| This study | Fecal sludge | 2380 mg/L | Total solids (pre-dewatering) |
| (Roldan-Hernandez et al. 2022) | Primary settled solids | 14-16% | Total solids (post dewatering) |
| (Mendoza Grijalva et al. 2022) | Wastewater influent | 900 mg/L | Total solids (post- dewatering |

Table S6. Linearized model fits for virus decay RNA

| **Target** | **Temperature** | **Intercept** | **Slope** | **R squared** | **p value** |
| --- | --- | --- | --- | --- | --- |
| N1 | 4 | -0.02853384 | 0.008996999 | 0.5150125 | 0.01945 |
| N1 | 15 | -0.59177872 | 0.008221262 | 0.1321031 | 0.271888 |
| N1 | 30 | 0.66077754 | -0.022970426 | 0.4695375 | 0.019966 |
| N2 | 4 | 0.19877029 | 0.010363543 | 0.4896914 | 0.02427 |
| N2 | 15 | 0.02755292 | 0.008322134 | 0.1484826 | 0.241869 |
| N2 | 30 | -0.29514539 | -0.022768482 | 0.5515161 | 0.008844 |
| PMMoV | 4 | 0.6884084 | 0.010987639 | 0.50482539 | 0.021287 |
| PMMoV | 15 | 0.8560139 | 0.009256381 | 0.62264304 | 0.003885 |
| PMMoV | 30 | 0.9862716 | 0.003002407 | 0.05852259 | 0.500709 |
| ORF12 | 4 | 0.348165 | 0.008144489 | 0.2487639 | 0.142258 |
| ORF12 | 15 | 0.3508941 | 0.006185107 | 0.3227463 | 0.068255 |
| ORF12 | 30 | 0.4171858 | -0.004595082 | 0.1086251 | 0.322287 |
| ToBRFV_Mo | 4 | 0.7515238 | 0.006959076 | 0.3027474 | 0.099355 |
| ToBRFV_Mo | 15 | 0.7903959 | 0.003879378 | 0.154808 | 0.231234 |
| ToBRFV_Mo | 30 | 0.9720335 | -0.012309629 | 0.41332 | 0.032874 |
